# A Multimodal Benchmark for Evaluating Cause-of-Death Inference Using Child Health and Mortality Data

**DOI:** 10.64898/2026.07.13.26357980

**Authors:** Junhe Yang, Soumyakanti Pan, Hyun Seung Lim, Yue Chu, Yuting Guo, Nishtha Agarwal, Varun Babbar, Gaurav Rajesh Parikh, Yiqun T. Chen, Chris A. Rees, Ziyaad Dangor, Sanjay G. Lala, Zehang Richard Li, Samuel J. Clark, Zhenke Wu, Abhirup Datta, Li Liu, Cynthia Rudin, Samuel V. Scarpino, Benjamin M. Gyori, Tyler H. Mccormick

**Author notes:** Joint first authors (Junhe Yang, Soumyakanti Pan).

## Abstract

Accurately attributing causes of death is vital for global health, yet fewer than 5% of deaths in resource-constrained regions are medically certified. To assign causes to these unlabeled deaths at scale, practitioners traditionally rely on verbal autopsy, using supervised statistical models to classify based on structured survey data. However, modern mortality surveillance increasingly collects rich, unstructured multimodal data, such as free-text caregiver narratives and postmortem diagnostics, which traditional supervised statistical models struggle to seamlessly integrate. In this paper, we present a comprehensive, multimodal benchmark for cause-of-death classification using data from the Child Health and Mortality Prevention Surveillance (CHAMPS) network, a unique surveillance platform spanning nine countries across South Asia and Sub-Saharan Africa. Using this dataset, we introduce an evaluation framework designed to rigorously assess diagnostic reasoning, moving beyond traditional metrics that fail to capture complex clinical realities. We demonstrate the utility of this benchmark by evaluating zero-shot large language models against supervised baselines across various data modalities. Our results reveal distinct differences in how these modeling approaches synthesize unstructured medical evidence. This benchmark provide a rigorously defined resource for assessing clinical reasoning in next-generation mortality surveillance.

## 1. Introduction

In low- and middle-income countries, infant and child mortality remains a critical public health crisis. While the staggering rates of these deaths are well-documented, the specific underlying causes often remain unrecorded (Horton, 2007; Hill et al., 2007; Setel et al., 2007). While identifying these causes is the foundational step toward targeted interventions, capturing the complex sequence of fatal events presents a important data synthesis challenge (Boerma and Stansfield, 2007). Historically, mortality surveillance in these resource-constrained settings has relied on verbal autopsy (VA) i.e., structured interviews conducted with surviving family members or caregivers after a death). However, as global health data infrastructure evolves, mortality investigations are expanding beyond simple surveys to include complex, multimodal data sources. Establishing rapid, automated, and highly accurate methods to synthesize this diverse data into definitive causes of death is essential for preventing future mortality, yet the machine learning community currently lacks robust benchmarks to evaluate predictive systems on this unique, high-stakes clinical task.

Automated cause-of-death classification has traditionally relied on statistical and machine learning models trained strictly on structured verbal autopsy data, e.g., InterVA (Byass et al., 2012), Tariff (James et al., 2011), InSilicoVA (McCormick et al., 2016), and others (WHO, 2022). While these approaches have shown promise, they are inherently bottlenecked by their dependence on standardized questionnaires, such as the World Health Organization (WHO) verbal autopsy instrument (World Health Organization, 2025). Existing automated VA frameworks mandate strict conformity to these rigid structures, which not only limits the incorporation of novel datasets but fundamentally restricts the integration of diverse, unstructured data modalities increasingly available in modern mortality surveillance. While recent work such as LAVA (Chen et al., 2025) has begun to demonstrate the potential of large language models (LLMs) for classification in this domain, current systems still largely under-utilize the rich contextual information contained in clinical narratives, laboratory diagnostics, and postmortem pathology. To bridge this gap, we introduce a novel benchmarking framework using multimodal data from the Child Health and Mortality Prevention Surveillance (CHAMPS) network. By aggregating demographic profiles, unstructured VA narratives, postmortem diagnostic results, and clinical abstraction records, we present the first multimodal predictive benchmark for CHAMPS data, positioning complex mortality classification as a rigorously evaluable machine learning task.

A central objective of this work is to advance the methods of evaluation in medical machine learning by comparing two fundamentally different paradigms for automated cause-of-death inference. The first paradigm comprises supervised statistical and machine learning models trained on gold-standard labels to predict mortality causes from structured inputs. The second leverages pretrained generative large language models (LLMs), which possess broad biomedical knowledge but have not been exposed to the labeled data. Rather than querying the models directly for a diagnosis, we construct structured prompts that present each case as a consolidated clinical dossier, framing the prediction as a constrained classification task refined through iterative prompt engineering and error analysis. Beyond simply benchmarking predictive performance, this comparison interrogates the evaluation methodology itself. We demonstrate that traditional exact-match classification metrics fail to capture the compounding clinical realities of mortality, necessitating a broader, set-based “causal-chain” evaluation framework, where the causal-chain comprises of a collection of causes that likely led to the death. By expanding the validation target to include any condition within the mortality sequence, we more accurately reflect clinical diagnostic logic. Ultimately, by stress-testing these models across progressive modality ablations, we provide a blueprint for evaluating the next generation of automated verbal autopsy systems, illustrating a transition from rigid questionnaires to flexible, multimodal clinical inference.

## 2. CHAMPS data

### 2.1. Data source

The Child Health and Mortality Prevention Surveillance (CHAMPS) network (https://champshealth.org/) is an ongoing, multi-country mortality surveillance platform operating in Bangladesh, Ethiopia, Kenya, Mali, Mozambique, Nigeria, Pakistan, Sierra Leone, and South Africa. In each participating country, CHAMPS investigators collaborate with community and public health officials to establish surveillance sites encompassing urban and rural settings with varying health system capacity, infectious disease burden, and sociodemographic characteristics. Each site maintains active mortality surveillance systems to rapidly identify stillbirths and deaths among children younger than five years of age. All deaths undergo standardized data collection and physician panel review to assign underlying and contributing causes of death (see Section 2.3).

### 2.2. Data modalities and variable domains

Each case in our benchmark is represented as a multimodal clinical dossier, synthesizing data from demographic surveillance, verbal autopsy, laboratory tests, and clinical abstractions. By bridging antemortem clinical trajectories and postmortem diagnostics, these modalities attempt to reconstruct the necessary evidence to evaluate complex reasoning across the sequence of fatal events.

#### 2.2.1. Basic demographics

In this article, we focus on the age group categories “Infant (28 days to *<*12 months)” and “Child (12 months to *<*60 months)”, corresponding to 2,191 cases^1^ from year 2016 through 2025. Neonatal deaths (including “Early Neonate (1–6 days)”, “Late Neonate (7–27 days)”, and “Death in the first 24 hours”) as well as stillbirths (“Stillbirth”) are excluded from the current analysis and are reserved for future investigation. Among the 2,191 cases, the raw dataset contains 26 basic demographic variables covering site identifiers, biological sex, hospitalization history, and anthropometric measurements (e.g., birth weight, limb measurements) that serve as indicators of nutritional status and developmental maturity. Overall, approximately 38% of values across these variables are missing, and two variables exhibit more than 90% missingness (see Figure 1). Unless otherwise specified, all subsequent analyses and summary statistics reported in this manuscript are restricted to these 2,191 cases in the infant and child cohort.

**Figure 1.**
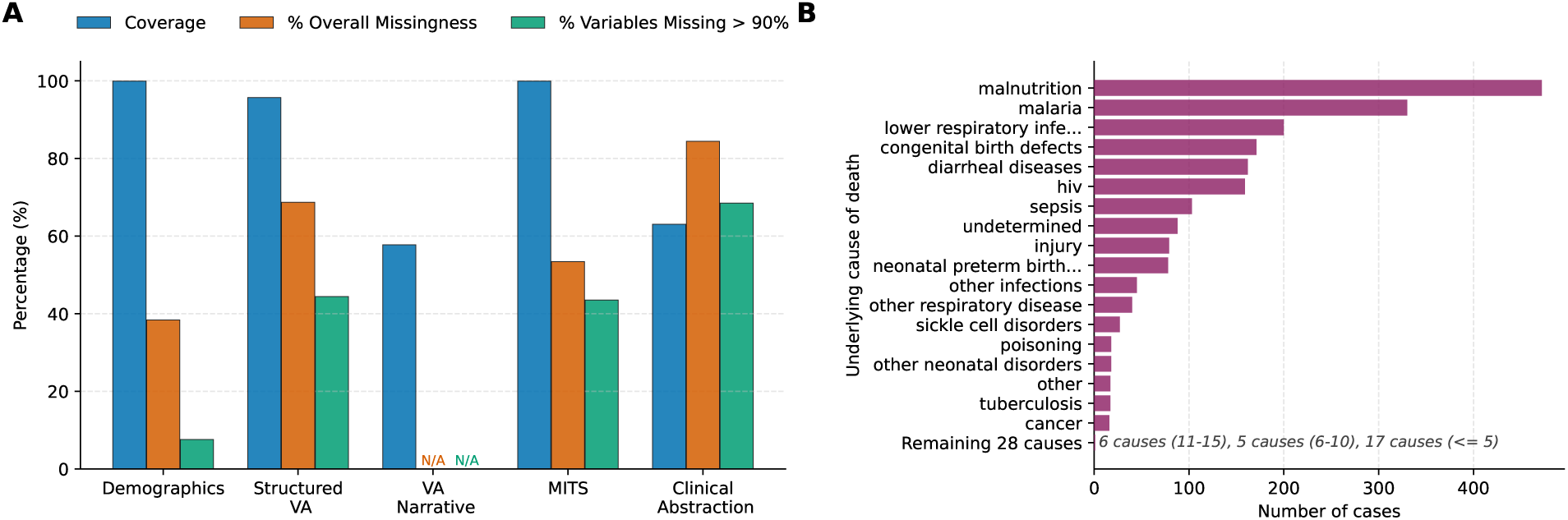
(A) Data quality metrics across data modalities in the CHAMPS infant and child cohort: missingness metrics are not defined for the VA narrative since it is a single unstructured text field. (B) Distribution of underlying causes of death: bars show the 18 causes out of 46 with ≥15 cases, highlighting the strong class imbalance in the dataset; among the remaining 28 causes, 6 causes have 11-15 cases, 5 causes have 6-10 cases, and 17 causes have less than or equal to 5 cases.

#### 2.2.2. Verbal autopsy

Verbal autopsy data are collected via standardized CHAMPS and Verbal Autopsy and Social Autopsy (VASA) instruments, comprising approximately 420 structured items covering symptom patterns, clinical history, and chronic conditions. Responses are typically recorded as binary indicators (“yes”/“no”/“don’t know”) or as duration measures. The structured instrument exhibits sparsity, with 68% overall missingness, and 45% of the items exceeding a 90% missingness rate. The structured VA instrument is followed by a de-identified free-text narrative in which the respondent is asked to describe, in their own words, the events that led to the child’s death, providing a chronological account of the child’s illness and care-seeking behavior. While the narrative field is missing for 42% of the cases, available entries are substantial, with a median word count of 121 (interquartile range: 432.7–971.2 characters). The narrative component offers complementary contextual information beyond the structured instrument, documenting nuanced symptom trajectories and barriers to care that escape predefined response fields.

#### 2.2.3. Child clinical abstraction

Clinical abstraction data are synthesized from health facility records to capture the child’s antemortem clinical course. These modules include medical history, immunization records, vital signs, and diagnostic testing for HIV and TB. The abstraction also contains a free-text case summary written in the style of a hospital discharge note, synthesizing clinical trajectories and pertinent diagnostic results. Only 63% of cases have associated clinical records, yielding a 723-variable feature set with 84% overall missingness. Approximately 69% of these variables exhibit missingness rates above 90% (Figure 1A).

#### 2.2.4. Minimally invasive tissue sampling (MITS)

The MITS procedure generates high-fidelity postmortem diagnostic data from blood, cerebrospinal fluid, and tissue specimens. Two major categories of diagnostic information that are derived from MITS investigations are – (1) TaqMan Array Card, providing pathogen detection across multiple gene targets, and (2) conventional microbiology and molecular assays, including Gram stains, cultures, and pathogen-specific PCR (e.g., HIV, malaria, and tuberculosis). Given the high dimensionality and sparse nature of these diagnostic assays, the combined MITS feature set exhibits substantial missingness. Across 525 selected variables, the overall missingness rate is 53%, with 44% of features exceeding 90% missingness (Figure 1A).

### 2.3. Cause of death

CHAMPS recognizes that death often results from a sequence of complex, interacting clinical conditions rather than a single isolated event. Accordingly, each case is represented by a “causal chain”, consisting of an underlying cause (the primary precipitating event), and a sequence of up to eight immediate and morbid conditions. These labels are ascertained by a local Determination of Cause of Death (DeCoDe) panel of experts (Blau et al., 2019) who synthesize all evidence from Section 2.2 to assign codes according to International Classification of Diseases (ICD) standards. These are subsequently mapped to the CHAMPS Basic U5 Mortality Categorization scheme.

Within the infant and child cohort, all cases include an underlying cause. However, label coverage for the remainder of the sequence varies: an immediate cause is recorded for 69% of cases, and a first additional morbidity is present in 39%. The distribution across 46 distinct underlying causes is markedly heterogeneous; while malnutrition (20%) and malaria (14%) are the most frequent, a long tail of 17 causes appears with five or fewer cases each (Figure 1B).

## 3. Methods

We consider the task of predicting causes of death using the multimodal data sources described in Section 2.2. Two distinct modeling paradigms are evaluated. First, we apply a range of statistical and machine learning methods, framing the problem as a supervised multi-label classification task in which models are trained on cases with gold-standard labels and evaluated on a held-out test set. Second, we use pre-trained generative large language models, prompting them with structured case dossiers to predict causes of death without access to the training labels. Across both paradigms, we systematically assess the contribution of individual data modalities and their combinations to predictive performance, as detailed in Section 4.

### 3.1. Statistical and machine learning models

For our supervised models, we evaluate three modeling paradigms: a domain-specific Bayesian hierarchical model, a gradient-boosting decision tree, and a regularized multinomial logistic regression. Tabular data was preprocessed directly, while unstructured narratives were converted into binary features by mapping text to ICD codes via a hybrid approach of generative LLMs and semantic embedding retrieval (see Section A1 in the Appendix).

#### 3.1.1. *InSilicoVA.* InSilicoVA

(McCormick et al., 2016) is a Bayesian hierarchical model that jointly estimates individual cause probabilities and population mortality fractions. To establish a baseline representative of traditional methods, we restrict its inputs to demographics and structured VA items, excluding narratives, MITS, and clinical abstractions. Continuous variables are discretized by quantiles, and VA responses are encoded as binary indicators or missing. The model treats the cause of death as a latent categorical variable, with features as conditionally independent “symptoms” governed by an empirical *probbase* matrix learned from a training data. We execute posterior inference via a Metropolis-within-Gibbs sampler using the InSilicoVA R package (Li et al., 2025), extending its internal utilities to support our custom training requirements.

#### 3.1.2. *LightGBM.* LightGBM

(Ke et al., 2017) is a gradient boosting framework designed for highdimensional data and mixed feature types. Unlike our InSilicoVA baseline, we utilize LightGBM to evaluate the full multimodal spectrum. A key architectural advantage is its native handling of missing data; the model treats missingness as an informative feature by learning optimal split directions. To address severe class imbalance, we apply balanced class weighting. Hyperparameters, including learning rate and tree depth, are optimized via 5-fold cross-validated grid search.

#### 3.1.3. Elastic Net

We evaluate a multinomial logistic regression with elastic net penalty (Zou and Hastie, 2005), combining *ℓ*_1_ and *ℓ*_2_ regularization for automatic feature selection and stability against collinearity. Unlike our other models, this framework requires complete numerical matrices. Continuous features underwent *k*-nearest neighbors imputation using KNNImputer from scikit-learn, and unit-variance scaling. Missing categorical values across VA, MITS, and clinical domains were zero-filled; while this conflates “unrecorded” with “negative” responses, it allows the model to operate without numerical distortion. We used the SAGA solver with balanced class weighting, optimizing the regularization strength and penalty ratio via 5-fold cross-validated grid search.

### 3.2. Generative large language models (LLMs)

In contrast to the supervised models in Section 3.1, our second modeling paradigm leverages pre-trained generative large language models (LLMs) to perform zero-shot classification. This approach completely forgoes model training; instead, the multimodal case data is transformed into structured clinical dossiers and passed directly to the model via guided prompts to predict the cause of death.

#### 3.2.1. Model selection

We evaluate two open-weight models from a consistent architectural lineage: gpt-oss-20b and gpt-oss-120b. Selecting a single model family minimizes confounding factors—such as disparate tokenization or instruction-tuning—enabling a controlled comparison of parameter scale (20B vs. 120B). Both models utilize the *Harmony* chat-oriented format (OpenAI, 2025), ensuring consistent prompt rendering and structured extraction. We prioritize these checkpoints under the Apache 2.0 license to facilitate community reproducibility. By holding input construction, prompt templates, and decoding configurations constant, we isolate the influence of model capacity on clinical reasoning. The 20B model serves as a computationally efficient baseline, while the 120B model is hypothesized to better synthesize long, heterogeneous evidence for fine-grained classification. This paired comparison provides an empirical assessment of how parameter scale impacts diagnostic accuracy and robustness under matched experimental conditions.

#### 3.2.2. Data transformation

To support zero-shot inference, we transformed the multimodal data into unified, analysis-ready text dossiers. Unlike numerical matrices, these dossiers provide semantic context by mapping raw field codes to human-readable descriptors using CHAMPS data dictionaries. For demographics and structured VA items, we explicitly excluded missing or non-informative responses (e.g., “don’t know”) to prevent the model from conditioning on noise. Valid categorical responses were normalized to canonical “yes”/“no” forms, while quantitative values (e.g., age, weight) were preserved. Clinical evidence streams, including MITS lab results, and clinical abstractions, were filtered and translated into clinically interpretable descriptive statements. This process yielded a consolidated per-case representation where all available modalities are organized into consistent sections. By synthesizing cleaned narrative text with semantically meaningful structured pairs and standardized clinical evidence, we provide a high-fidelity input for downstream prompting. For evaluation, reference causes of death were harmonized to a static label space.

#### 3.2.3. Prompt design

To ensure comparability across model scales, we established a fixed prompting protocol using the *Harmony* chat-completion format, holding section ordering, system instructions, and decoding hyperparameters strictly constant. Each case was formatted as a structured dossier with modular sections corresponding to distinct clinical modalities (Section 2.2), which could be included independently or in combinations as human-readable statements. This architecture provides a stable input format for heterogeneous records while mitigating the influence of missing entries. We framed inference as a constrained classification task, providing the allowed label schema, and instructing the model to select predictions exclusively from this list. To facilitate automated parsing, we enforced a strict JSON output schema requiring the top three candidate causes ranked by confidence, using exact label strings to avoid normalization errors. This standardized design constrains the prediction space and enforces a structured format suitable for consistent downstream evaluation.

#### 3.2.4. Prompt engineering

Beyond the structural design, the prompt’s clinical guidance underwent an extensive, iterative engineering process. This guidance was developed through rigorous error analysis of pilot runs. We examined confusion matrices and manually reviewed model-generated predictions to identify recurring misclassifications and ambiguous decision boundaries within the target label schema. These observed failure modes were then translated into targeted, label-specific diagnostic heuristics. Candidate guidance statements were drafted and refined through a combination of LLM-assisted generation and expert human adjudication. Once finalized, these refined instructions were incorporated into the system prompt to mitigate the most frequent and consequential errors, while ensuring the overarching prompting protocol remained strictly fixed across tested models and evaluation regimes. The complete engineered guidance is provided in Section A2 of the Appendix.

## 4. Evaluation

### 4.0.1. Train-test split

We established a universal split to ensure comparability across different modeling paradigms. The test set prioritizes unstructured text by filtering the 58% of the cohort with narratives (minimum 30 characters). We allocated 20% of this subset (252 cases; ∼12% of the total cohort) as the held-out test set, with the remaining 1,939 cases—including all instances without narratives—forming the training set. Stratified random sampling on the underlying cause ensures the test set mirrors the cohort’s label distribution, enabling a direct evaluation of zero-shot LLMs against supervised models on identical, narrative-rich cases.

### 4.0.2. Data domain ablation design

To identify the minimal evidence threshold required for robust mortality classification, we progressively expand input modalities across five phases that mirror the logistical and financial constraints of global health surveillance. We begin with community-level data (Phases 1–2: *Structured VA* and *demographics*), then integrate *VA narratives* (Phase 3) to assess signal extraction from raw text, and finally incorporate high-fidelity facility diagnostics (Phases 4–5: *MITS* and *clinical abstractions*). This additive approach allows us to quantify the marginal utility of specialized clinical evidence relative to its collection cost, effectively simulating the trade-offs between diagnostic accuracy and resource availability in real-world settings.

### 4.1. Evaluation metrics

Because cause-of-death assignment involves complex, interacting clinical sequences (Section 2.3), we systematically evaluate model performance using set-based agreement. For any case, we define a successful prediction by the intersection between a generated prediction set P and a ground-truth validation set V, i.e., a prediction is successful if P ∩ V ̸= ∅□. To comprehensively capture model performance, we manipulate the cardinalities and compositions of P and V to establish the following evaluation frameworks. For the prediction set P, we evaluated performance at two cardinalities: top-1 (|P| = 1) and top-*k* (|P| = *k*), where *k* = 3. For supervised models, P contains the causes with the *k* highest output probabilities. For zero-shot generative LLMs, P was generated via explicit prompting, instructing the model to: *“Pick the top 3 most likely causes of death from the allowed causes only, ranked by probability (highest first)”* (see Section A2 in the Appendix for details).

#### 4.1.1. Matching criteria

Recognizing the complexity of the CHAMPS causal chain (Section 2.3), we defined two evaluation regimes based on the composition of the validation set V:

- *Single Cause Evaluation:* The validation set V is restricted to the underlying cause of death. For supervised models, this mirrors the standard classification setup where training and evaluation labels are identical. This metric assesses the model’s ability to isolate the primary precipitating event leading to the fatality.
- *Anywhere-in-Chain evaluation:* The validation set V is expanded to include the entire causal chain, encompassing the underlying, immediate, and all contributing comorbid conditions. For supervised models, this introduces a clinically-informed “fuzzy” evaluation, i.e., while models are trained strictly on the underlying cause, test predictions are validated against any condition in the full chain. For zero-shot LLMs, this simply expands the target label set. This logic acknowledges clinical reality; for example, if a model identifies a fatal condition such as “sepsis,” it should not be penalized if the expert panel categorized it as an immediate rather than underlying cause.

#### 4.1.2. Individual-level metrics

Based on the intersection rule (P ∩ V ̸= ∅□), our primary metric is overall accuracy, the proportion of test cases with a successful prediction. We report four formulations: *Top-1* and *Top-3 Single Cause accuracy*, validated against the underlying cause, and *Top-1* and *Top-3 Anywhere-in-Chain accuracy*, validated against the full clinical causal chain. This evaluates performance across both diagnostic confidence (|P| ∈ {1, 3}) and clinical breadth.

#### 4.1.3. Class-specific metrics

Beyond overall accuracy, we compute precision and recall for each cause-of-death category under both evaluation regimes. To operationalize these via set-based matching, a prediction *c* ∈ P is a true positive for class *c* if *c* ∈ V, and a false positive if *c* ∈*/* V. Conversely, a reference cause *c* ∈ V is a false negative if *c* ∈*/* P. We then calculate class-wise *precision* (ratio of true positive and all positive predictions for a class) and *recall* (ratio of true positives and all actual instances of a class) for each cause-of-death category.

#### 4.1.4. Population-level metrics

To evaluate aggregate estimation, we calculate the *Cause-Specific Mortality Fraction* (CSMF) accuracy. Let V^uc^ denote the singleton set containing the underlying cause for case *i*. We define the true CSMF for cause-of-death *c* by 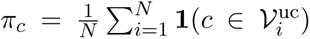,

where **1**(·) denotes the indicator function, and *N* is the total number of cases. Moreover, define the predicted CSMF for cause-of-death *c* by 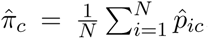, where 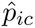 denotes the predicted probability of cause *c* for case *i*, as estimated by a model. For supervised models, obtaining 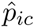 is straightforward as they usually return predicted probabilities across the entire label schema. For LLMs, we reconstruct these values using the reported probabilities for the top-3 causes and assign zero to all remaining causes. Following Murray et al. (2011), we have

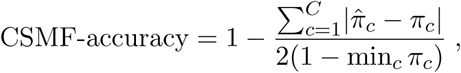

where *C* corresponds to the total number of cause-of-death categories. This metric measures the proximity of estimated cause fractions to the truth, ensuring reliable mortality surveillance for public health decision-making despite individual-level uncertainty.

## 5. Results

We benchmark zero-shot LLMs against fully supervised models trained on the CHAMPS cohort across four dimensions: predictive accuracy, cumulative clinical evidence ablation, cause-specific performance, and population-level mortality distributions. Appendix A3 provides additional evaluation results beyond the following metrics.

### 5.1. Overall accuracy

Supervised models attained the highest accuracy; LightGBM peaked at 59.5% Top-1 (77.8% Top-3) under the Single Cause regime, while the top zero-shot LLM, gptoss-120b, reached 48.8% Top-1 (68.3% Top-3). However, evaluation under the Anywhere-in-Chain regime significantly shifted these dynamics, disproportionately benefiting LLMs (Figure 2). In this setting, the gap between LightGBM (68.2% Top-1) and gpt-oss-120b halved, narrowing from an 11-point deficit to less than 5 points.

**Figure 2.**
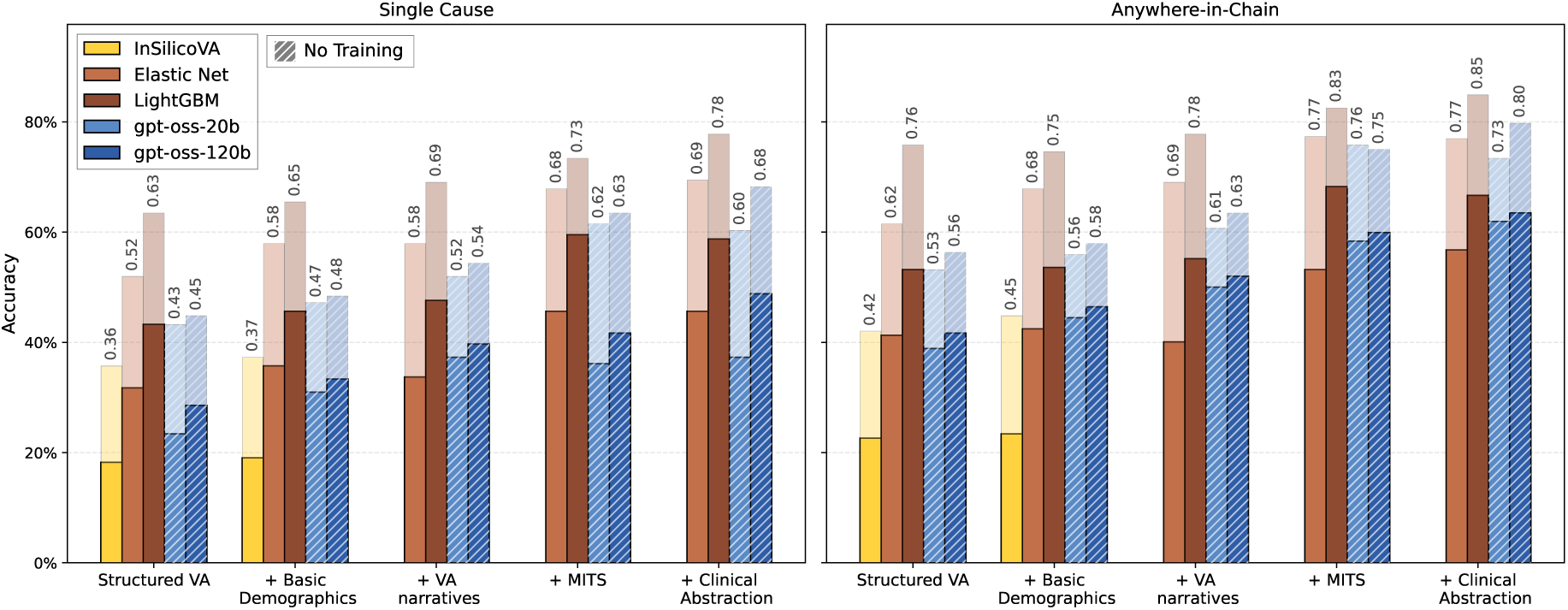
Predictive performance across additive clinical data ablations. Left and right panels display accuracy under the Single Cause and Anywhere-in-Chain evaluation regimes, respectively. The x-axis represents the stepwise addition of data modalities, progressing from structured VA alone to the full clinical dossier. For each data stage, the primary solid bar denotes Top-1 accuracy, while the overlaid extended bar indicates Top-3 accuracy.

Beyond raw accuracy, the paradigms exhibited divergent scaling as clinical evidence accumulated. Supervised models established high baselines via structured data but plateaued as narratives and MITS were added. Conversely, LLMs leveraged this complexity to drive a nearly 22-point Top- 1 surge. This suggests supervised models optimize for structured targets, whereas LLMs better synthesize heterogeneous clinical evidence into a comprehensive picture of physiological collapse.

### 5.2. Impact of cumulative clinical evidence

Stepwise data ablation highlights divergent learning curves between paradigms. Restricted to structured verbal autopsy (VA), LLMs exhibited a significant performance deficit; gpt-oss-120b trailed LightGBM by 15 points in Top-1 Single Cause accuracy (28.6% vs. 43.3%). Narrative integration marked a critical inflection point: while LightGBM gained only 2 points from text, gpt-oss-120b saw a 6.4-point Top-1 surge and a 5.6-point gain in Top-3 Anywhere-in-Chain accuracy. These narratives allowed LLMs to untangle complex symptom profiles that structured surveys often obscure.

Objective MITS data improved broad candidate retrieval but did not uniformly enhance Top-1 discrimination. While MITS drove 9-point Top-3 gains for both LLMs, Top-1 performance remained muted, with gpt-oss-20b even declining slightly from 37.3% to 36.1%. This suggests that while highfidelity postmortem findings serve as definitive diagnostic anchors that pull the correct cause into the top three candidates, they also introduce specific clinical noise that can disrupt precise Top-1 ranking. Finally, clinical abstractions yielded peak performance under broader evaluation despite data sparsity. In the Anywhere-in-Chain regime, LightGBM and gpt-oss-120b reached overall Top-3 peaks of 84.9% and 79.8%, respectively. The 120b model consistently navigated this multimodal complexity better than its 20b counterpart, confirming that parameter scale provides a distinct advantage when reasoning over increasingly dense clinical contexts.

### 5.3. Cause-specific performance

Focusing on the complete clinical configuration, we compare class-wise precision and recall for our top-performing models, LightGBM and gpt-oss-120b (see Figure 3). This reveals a trade-off between the precision-oriented supervised paradigm and high-recall zero-shot reasoning. LightGBM shows superior precision in malnutrition (0.89 vs. 0.60 for gpt-oss-120b), leveraging training support. Conversely, gpt-oss-120b yields higher recall for lower respiratory infections (0.81 vs. 0.48). This suggests supervised models optimize for specificity, whereas LLMs act as broader screeners, capturing more true cases by synthesizing overlapping symptoms that calibrated classifiers might exclude.

**Figure 3.**
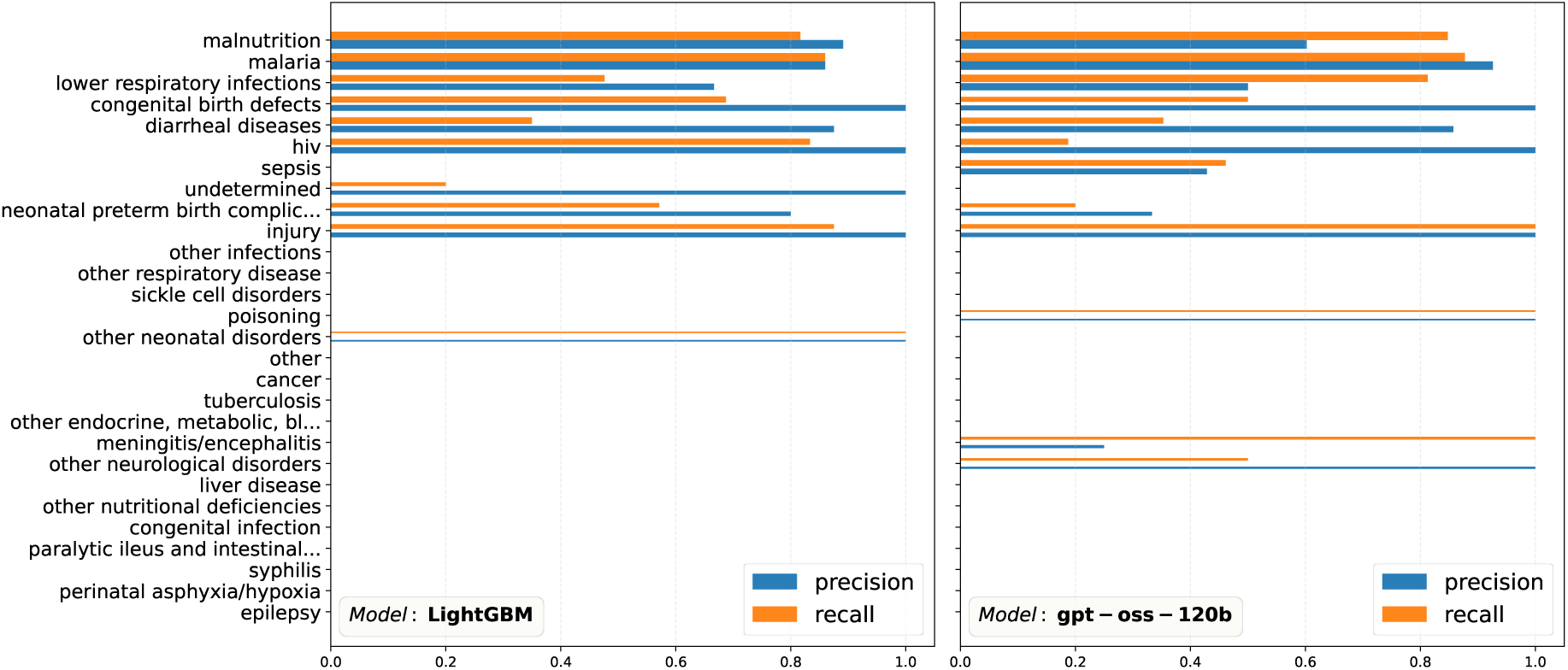
Class-wise performance comparison: Precision and recall under the Anywhere-in-Chain regime for LightGBM (left) and gpt-oss-120b (right) across 28 CHAMPS cause-of-death labels. Causes are ordered by training set frequency (top to bottom). Bar thickness scales with test-set support; thinner lines denote higher evaluation uncertainty for rare categories.

Malaria emerges as a notable point of convergence; remarkably, zero-shot gpt-oss-120b (0.93 precision, 0.88 recall) marginally outperforms LightGBM (0.86 precision, 0.86 recall). This indicates that for well-characterized infectious diseases with distinct clinical presentations, pre-trained reasoning can reach or exceed the performance of models trained on domain-specific cohorts.

### 5.4. Population-level mortality distributions

Aggregate CSMF accuracy did not follow a monotonic relationship with data volume. LightGBM reached peak fidelity (0.90) using only structured VA, yet both supervised models and LLMs experienced declining accuracy after introducing MITS diagnostics: gpt-oss-120b dropped from 0.69 to 0.59, and LightGBM from 0.88 to 0.86 (Table 1). This suggests high-fidelity postmortem evidence improves individual-level retrieval but may introduce distributional biases that skew aggregate fractions. Elastic Net was the exception, scaling consistently to a peak of 0.80 with the full dossier. These results underscore a nuanced trade-off in mortality surveillance: models optimized for individual-level accuracy do not always yield the most reliable estimates of population-level disease burden.

**Table 1.** Cause-specific mortality fraction (CSMF) accuracy across models and data modalities. S: Structured Verbal Autopsy, D: Basic Demographics, N: Verbal Autopsy Narratives, M: Minimally Invasive Tissue Sampling, C: Child Clinical Abstraction. Ticks imply inclusion of that data modality in the model.

| Data Configuration |  |  |  |  | CSMF Accuracy |  |  |  |  |
| --- | --- | --- | --- | --- | --- | --- | --- | --- | --- |
| S | D | N | M | C | InSilicoVA | Elastic Net | LightGBM | gpt-oss-20b | gpt-oss-120b |
| ✓ | ✗ | ✗ | ✗ | ✗ | 0.534 | 0.741 | 0.905 | 0.593 | 0.633 |
| ✓ | ✓ | ✗ | ✗ | ✗ | 0.523 | 0.716 | 0.877 | 0.674 | 0.679 |
| ✓ | ✓ | ✓ | ✗ | ✗ | — | 0.723 | 0.879 | 0.706 | 0.686 |
| ✓ | ✓ | ✓ | ✓ | ✗ | — | 0.765 | 0.859 | 0.592 | 0.593 |
| ✓ | ✓ | ✓ | ✓ | ✓ | — | 0.804 | 0.864 | 0.603 | 0.625 |

## 6. Discussion

### 6.1. Synthesis of modeling paradigms

We evaluated supervised classifiers against zero-shot LLMs for cause-of-death assignment across five clinical data configurations and two validation regimes. Importantly, this work does not seek to establish a direct performance competition between supervised statistical learning and zero-shot generative models. Given the fundamental differences in their training objectives, discriminative for the former and generative for the latter, we emphasize that comparing their raw metrics is an exercise in understanding trade-offs rather than identifying a superior model class. Our results indicate that these paradigms are functionally distinct, i.e., while supervised models are optimized for population-level point estimation, LLMs demonstrate a superior capacity for synthesizing evidence from unstructured narratives, particularly when evaluated under the “anywhere-in-chain” regime which accounts for complex co-morbidities.

### 6.2. Limitations and intended use

As detailed in our results, while larger model size and multimodal inputs yield significant gains for common infectious and injury-related causes, performance remains persistently low for rarer or clinically overlapping categories, such as HIV and Tuberculosis. Furthermore, while we establish the efficacy of zero-shot “off-the-shelf” LLMs, we do not explore domain-specific fine-tuning in this paper. We intentionally prioritize the zero-shot paradigm to simulate real-world feasibility in resource-constrained settings where the specialized technical expertise or computational infrastructure required for bespoke model training may be unavailable. To comply with the CHAMPS Data Transfer Agreement (DTA), we utilize locally hosted open-source models instead of commercial APIs, ensuring data remains within our secure internal architecture. Additionally, while our benchmark represents a diverse multi-country cohort, it is primarily focused on pediatric mortality (ages 0–5), not including stillbirths and neonatal deaths, and hence, the findings may not generalize to adult populations. The caregiver narratives are also subject to cultural and linguistic nuances that may affect model performance across different sites. Finally, our “anywhere-in-chain” evaluation relies on expert-grounded labels which, while rigorous, are subject to the inherent uncertainties of clinical cause-of-death determination in settings without gold-standard pathology.

### 6.3. Ethics and governance

The primary data utilized in this benchmark were collected by the CHAMPS network under protocols approved by the institutional review boards and national ethics committees at each surveillance site, with informed consent obtained from the parents or guardians of all children included. This specific study involves the secondary analysis of de-identified data; as the data pertains to deceased individuals and is anonymized, it does not constitute human subjects research under standard regulatory frameworks. Nonetheless, this work was conducted in accordance with the ethical standards of our institution and the strict terms of the CHAMPS Data Use Agreement (DUA), which limits usage to public health research and prohibits any attempts at re-identification.

### 6.4. Future work

Moving ahead, we will focus on bridging the identified performance gaps through agentic frameworks (Park et al., 2023) designed to emulate the collaborative adjudication of clinical panels. This includes implementing Tree-of-Thought (Yao et al., 2023) reasoning to traverse the hierarchical ICD-11 taxonomy. Finally, improving the calibration of LLM output probabilities remains a priority (see e.g., Shen et al., 2024) to ensure that generative models can produce reliable evidence for public health decision-making.

## Code availability and data access protocol

The complete suite of resources for this benchmark is hosted across three specialized anonymous repositories.The implementation of the supervised statistical and machine learning models, along with a pipeline to pre-process raw CHAMPS data, is provided at https://anonymous.4open.science/r/champs_statsML-F544/, the LLM-based pipeline is located at https://anonymous.4open.science/r/champs-llm-4E5D/, and the expert-in-the-loop annotation pipeline documentation is available at https://anonymous.4open.science/r/icd10-grounder-code-08EE/.

Due to the sensitive nature of the Child Health and Mortality Prevention Surveillance (CHAMPS) data, which involves clinical narratives and postmortem diagnostics from vulnerable populations, the full dataset is subject to a Data User Agreement (DUA) and cannot be hosted in public repositories. Researchers seeking the full CHAMPS Level 2 de-identified dataset for replication or extension can request access via the formal CHAMPS data portal (https://champshealth.org/causes-of-death/). Upon gaining access, our provided preprocessing scripts can be used to reconstruct the exact training and testing partitions described in this paper. The train-test split required to replicate the findings of this paper can be found in https://anonymous.4open.science/r/champs_statsML-F544/data/infant_child_split.json.

## Data Availability

Due to the sensitive nature of the Child Health and Mortality Prevention Surveillance (CHAMPS) data, which involves clinical narratives and postmortem diagnostics from vulnerable populations, the full dataset is subject to a Data User Agreement (DUA) and cannot be hosted in public repositories. Researchers seeking the full CHAMPS Level 2 de-identified dataset for replication or extension can request access via the formal CHAMPS data portal (https://champshealth.org/causes-of-death).

## Acknowledgement

This work used computational resources and storage services of the Hyak Klone and Tillicum clusters provided by the University of Washington and the eScience Institute.

## Appendix A1. Automated annotation of VA narratives

To extract medically salient concept mentions from free-text narratives and ground them to ICD- 10 codes, we developed a retrieval-augmented generation (RAG)-based grounding pipeline, inspired by the retrieval-augmented medical coding framework proposed by Baksi et al. (2025). The pipeline operates in three stages.

First, a generative large language model (LLM) performs structured concept extraction: given the input text, the model identifies all disease or clinical concepts mentioned and returns, for each, the verbatim text spans from the input that support it. Second, each extracted concept is combined with its supporting evidence into a single query string, encoded using the all-MiniLM-L6-v2 sentence transformer, and used to retrieve the top-*k* most semantically similar candidate terms from a prebuilt embedding index of ICD-10 codes. Third, an LLM call re-ranks the retrieved candidates for each concept in context, considering concept alignment, evidence alignment, and term specificity. To prevent hallucinations in the re-ranking step, the LLM is instructed to return term identifiers from the provided candidate list; any identifier returned by the LLM that does not match a retrieved term is discarded, ensuring final grounding is always drawn from retrieval set.

To produce character-level offsets for each grounded mention, the LLM-extracted evidence spans are mapped back to the original input text via a two-pass algorithm combining exact substring matching and fuzzy token-window sliding. This yields precise start/end character offsets for each evidence span in the source text. We applied this pipeline (using GPT-4o-mini as the LLM) to the CHAMPS de-identified verbal autopsy dataset, filtering on the open-ended narrative field and processing 4,009 non-empty records. Each output record contains the extracted disease concepts, their character-level evidence spans in the original narrative, and a ranked list of ICD-10 code assignments with similarity scores.

**Figure A1.**
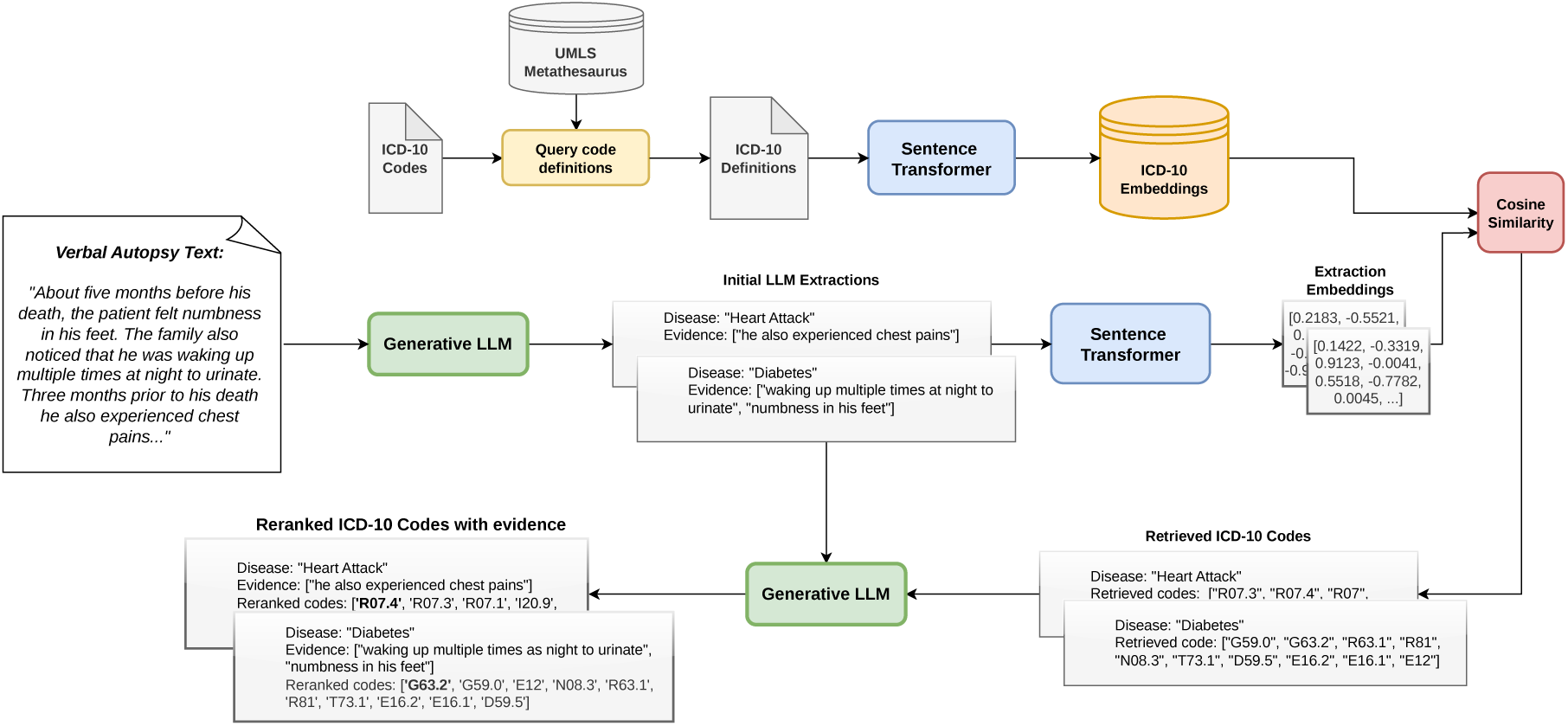
Pipeline for annotating narratives with ICD-10 codes

## Appendix A2. Details of the LLM-based approach

The final prompt template is the result of an iterative refinement process involving clinical experts and error analysis on a small pilot subset of the CHAMPS cohort. These early iterations focused on simple zero-shot classification without specific guidance, which led to several systematic failure modes. The inclusion of the GUIDANCE section was critical for steering the model toward clinical logic consistent with mortality surveillance standards.

### A2.1. Details of iterative refinement

During development, we identified the following areas where unguided LLMs diverged from expert diagnostic standards. We elaborate as follows

- *Proximate vs. underlying conflict (Malnutrition):* LLMs frequently prioritized proximate infections (e.g., lower respiratory infection/LRI or Diarrhea) over severe underlying malnutrition, even when clinical indicators like MUAC < 11.5 cm were present. To resolve this, we added explicit priority logic stating that Malnutrition should be the Top-1 cause in the presence of severe wasting, reflecting the hierarchical nature of global health mortality classification.
- *“Sepsis” bias:* Without specific boundaries, LLMs tended to over-utilize Sepsis as a default label for any systemic illness. The guidance was refined to force a distinction between Sepsis and Lower respiratory infections by requiring models to look for primary respiratory foci (chest indrawing, fast breathing) before defaulting to a systemic diagnosis.
- *Diagnostic over-reach (Malaria and HIV):* Initial prompts often led models to select Malaria based on fever alone or HIV based on vague maternal history. We implemented strict diagnostic thresholds requiring lab-confirmed results or advanced clinical markers (e.g., specific HIV testing or malaria TAC results) to prevent the inflation of these causes based on general geographic prevalence rather than case-specific evidence.
- *Neonatal Distinction:* Early prompts struggled with the nuance of neonatal mortality, often conflating prematurity with infection. Explicit guidance was added to separate Neonatal preterm birth complications from Neonatal sepsis based on the presence or absence of inflammatory signs, mirroring the DeCoDE panel’s adjudication logic.

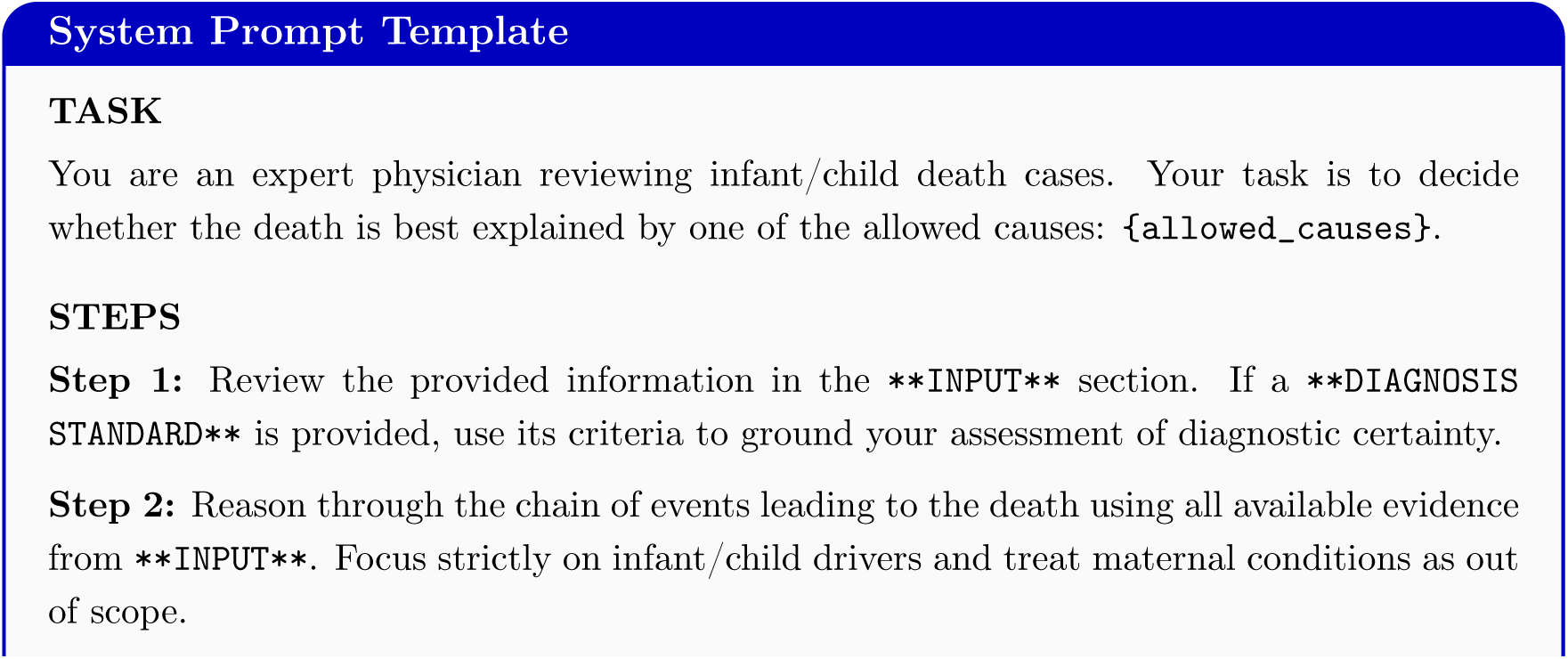
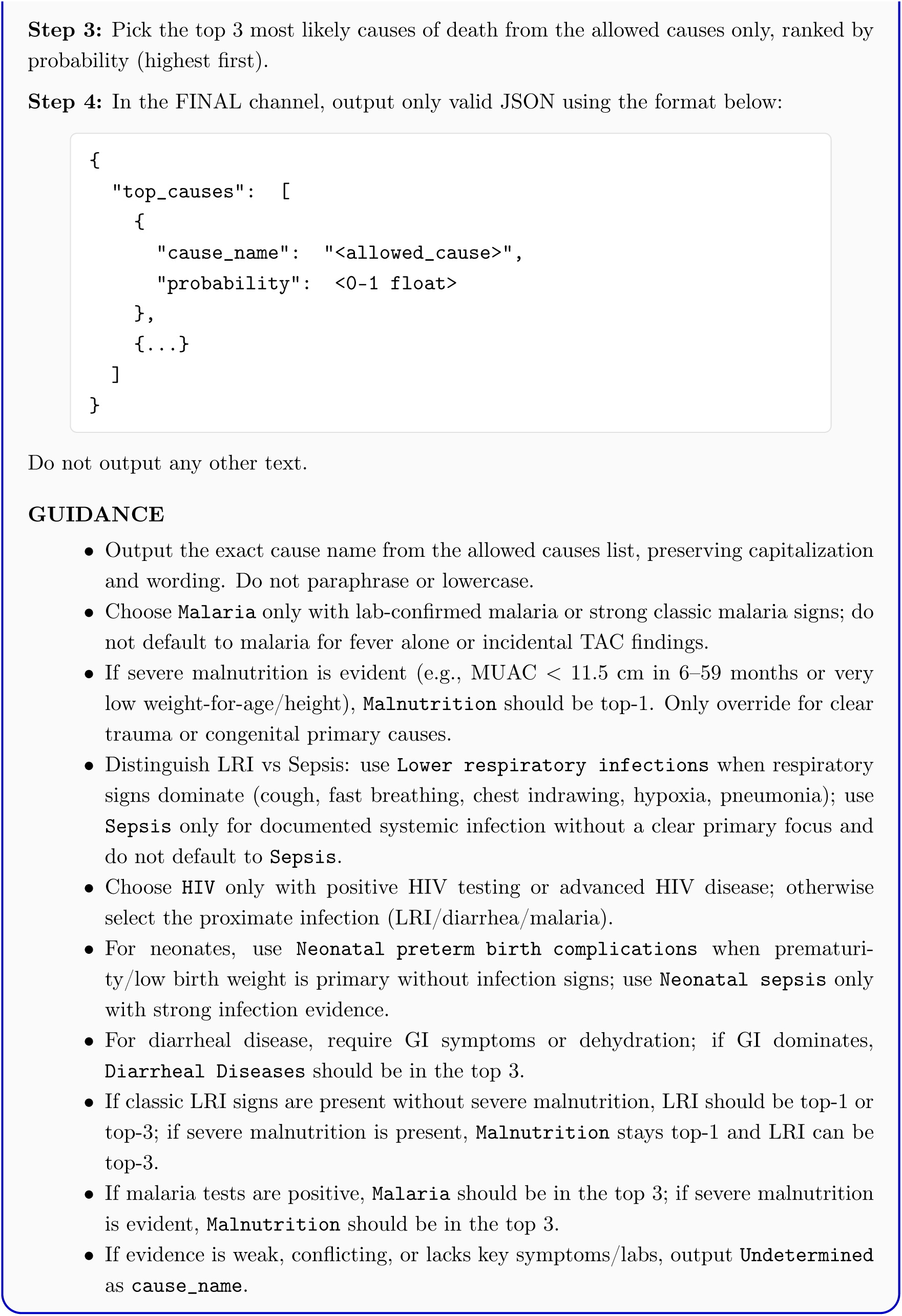

## Appendix A3. Additional results

### A3.1. Comparison of F1 scores

#### A3.1.1. Micro-F1 and evaluation regimes

The transition from Single Cause to Anywhere-in-Chain evaluation yields substantial performance gains across all models, with Micro-F1 scores, which reflects overall predictive accuracy, consistently increasing by approximately 10 to 15 percentage points (see Figure A2). LightGBM demonstrates the highest overall performance, peaking at a Micro-F1 of 0.68 under the Anywhere-in-Chain regime when utilizing the full multimodal feature set. The large language models exhibit a strong positive response to unstructured data; notably, gpt-oss-120b achieves a Micro-F1 of 0.63 in the full-input Anywhere-in-Chain configuration, significantly narrowing the gap with the strongest supervised baseline. Across all architectures, the integration of demographics, narratives, and clinical abstractions follows a monotonic improvement trend, confirming that multimodal evidence is essential for capturing the diagnostic breadth of complex mortality events.

#### A3.1.2. InSilicoVA and macro-level class balance

Macro-F1 scores, which provide an unweighted average across all 28 cause-of-death labels, highlight a distinct performance characteristic of the domain-specific InSilicoVA model (see Figure A3). In structured-only VA settings, InSilicoVA achieves a high Macro-F1 (0.24 for Single Cause), nearly doubling the performance of generic discriminative classifiers like LightGBM (0.12) and Elastic Net (0.13). Notably, InSilicoVA is the only model whose Macro-F1 exceeds its Micro-F1 in these settings, suggesting that its probabilistic architecture, which simultaneously target both individual and population-level accuracy, achieves a superior class-wise balance by avoiding the majority-class bias inherent in generic machine learning and zero-shot large language models. While these generic models tend to optimize for high-frequency categories, InSilicoVA maintains a more equitable sensitivity across the broad label space when limited to structured survey data.

**Figure A2.**
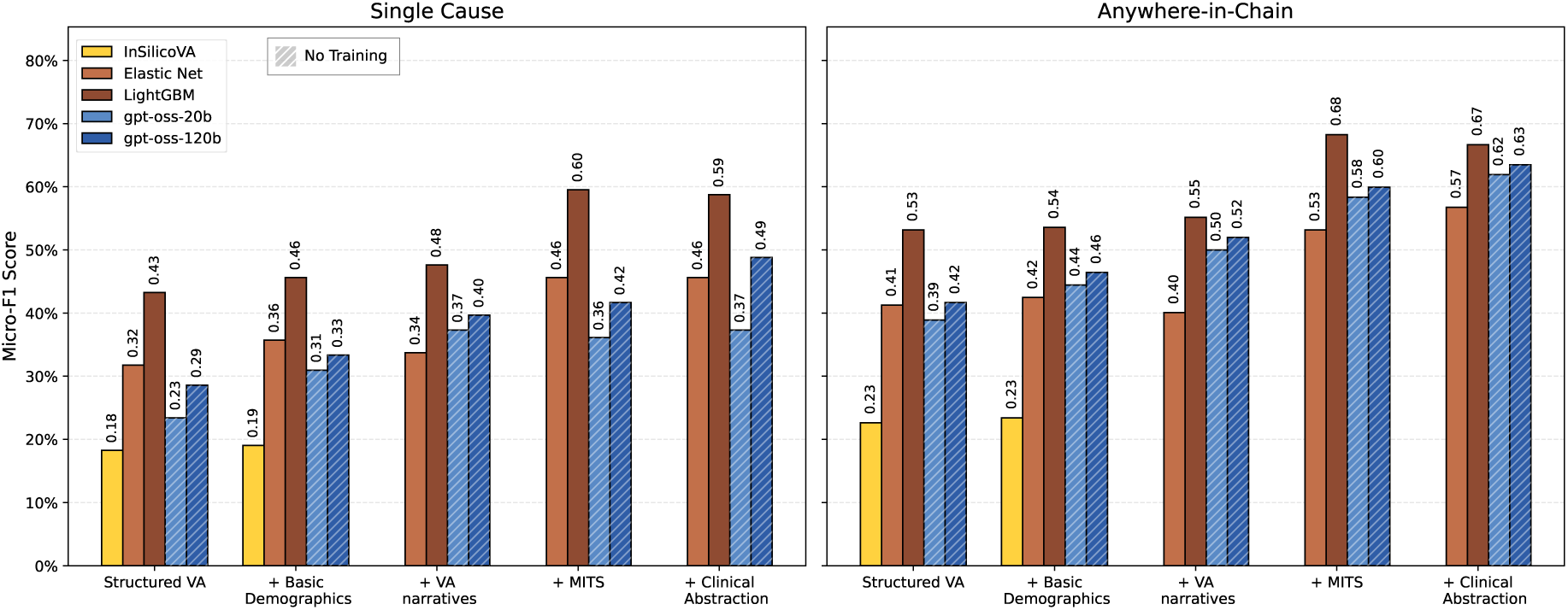
Micro-F1 scores across incremental clinical data ablations: Left and right panels display performance under the Single Cause and Anywhere-in-Chain evaluation regimes, respectively. Bars represent the Micro-F1 score for each method across the stepwise addition of data modalities.

**Figure A3.**
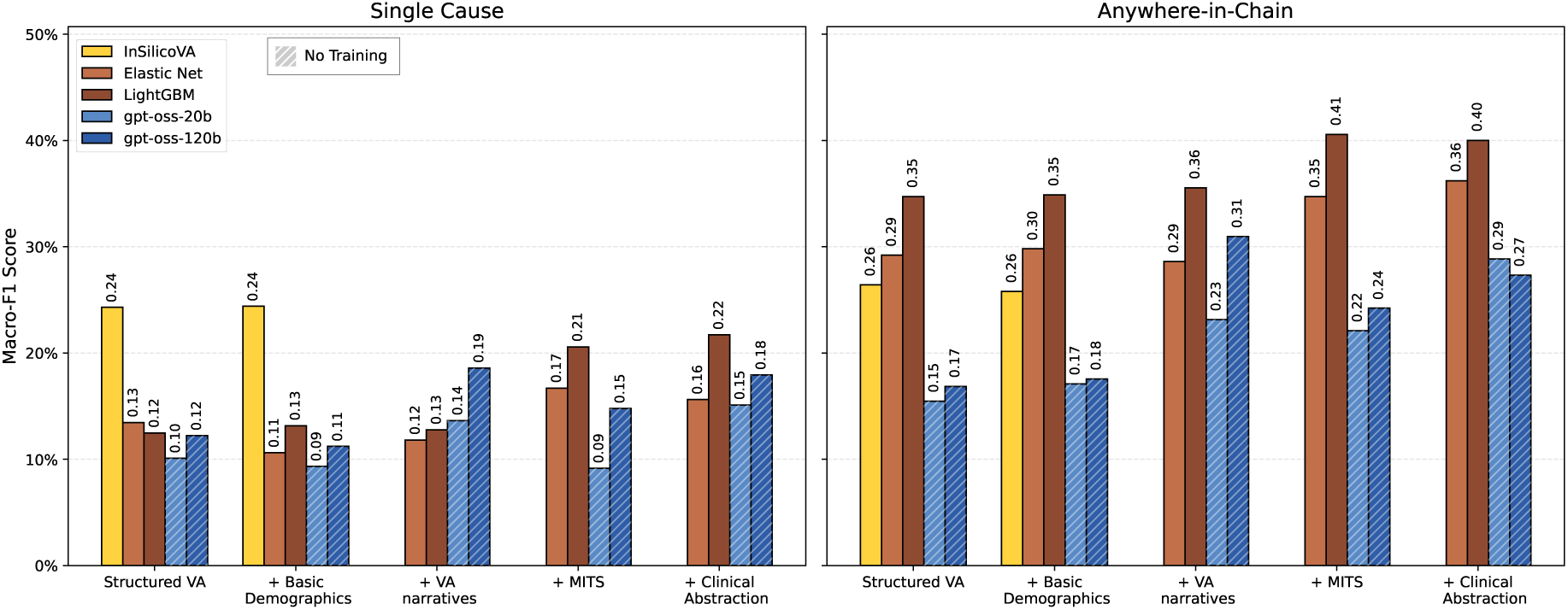
Macro-F1 scores across incremental clinical data ablations: Panels compare performance under Single Cause (left) and Anywhere-in-Chain (right) regimes. Bars denote macro-averaged F1 scores, providing an unweighted measure of predictive performance across all cause-of-death categories.

#### A3.1.3. Multimodal Impact on Rare Causes

The introduction of unstructured narratives and clinical abstractions acts as the primary driver for performance gains in rare or clinically overlapping categories. For gpt-oss-120b, the inclusion of verbal autopsy narratives nearly doubles the MacroF1 under the Anywhere-in-Chain regime, increasing it from 0.18 to 0.31. This suggests that while supervised models like LightGBM excel at population-level point estimation for common causes, LLMs provide superior semantic reasoning required to resolve the “long-tail” of the cause-of-death distribution. Our results demonstrate that peak macro-level performance is contingent upon the integration of multimodal evidence. The synergy between structured indicators and unstructured narratives suggests that resolving complex diagnostic categories requires a departure from traditional structured surveys, which often lacks the granular context necessary to distinguish between clinically overlapping causes of death.

## Footnotes

1 Data cutoff date: February 01, 2026

